# Geotemporal analysis of COVID-19 in the Dominican Republic 2020-2021

**DOI:** 10.1101/2023.01.27.23285032

**Authors:** Andreina Moreno, Carla Gonzalez, Lilian Pimentel, Demián Herrera, Manuel Colomé

## Abstract

**Introduction:** Coronavirus disease (COVID-19) was first identified in China in December of 2019 and has spread globally since. The Dominican Republic confirmed its first case on March 1^st^, 2020.

**Objective:** To analyze the spatial distribution of the incidence of COVID-19 and its correlation with the Human Development Index in the Dominican Republic from March of 2020 to March of 2021.

**Methods:** The cumulative incidence rates of COVID-19, number of deaths, lethality, mortality and Human Development Index of the provinces in the Dominican Republic were used from governmental sources and were analyzed in the Microsoft Excel 2016 program.

**Results:** Duarte was the province with the highest mortality per 100.000 inhabitants (68.94), followed by the Distrito Nacional (71,613 cases), Santo Domingo (49,759 cases) and Santiago (27,632 cases) with the highest number of cases. The 7-day moving average peak for new cases was July 30 of 2020 and the peak for new deaths occurred on September 6 of 2020. The highest positivity rate (40%) was reported in August of 2020. Lastly, an increase of 1.0 on the Human Development Index corresponded to a 10.7% increase in the incidence rates per province.

**Conclusion:** Prevention strategies should be strengthened by the Dominican government to reduce the contagion curve and thus reduce its spread and impact on the Human Development Index

## Introduction

Coronavirus disease 2019 (COVID-19) was first identified in December 2019 in Wuhan, the capital of the Chinese province of Hubei^1^. Since then, it spread globally and declared a pandemic four months later. Nowadays, coronavirus disease 2019 poses a significant threat to global health. The World Health Organization (WHO) declared this outbreak a “public health concern of international relevance” in January 2020. ^2^ COVID-19 has caused a total of 120,915,219 confirmed cases and 2,674,078 deaths globally, reporting 248,502 registered cases and 3,262 deaths in the Dominican Republic (DR) by March 2021.^3^ The SARS-CoV-2 is mainly transmitted through respiratory droplets during close contact with an infected individual. The infection can be transmitted by asymptomatic, presymptomatic, and symptomatic carriers. Amongst the most common symptoms include fever, dry cough and shortness of breath. ^3,4^ The first case confirmed by the DR was on March 1, 2020 imported by an Italian tourist and has since been expanded nationwide.

The epidemiological dynamics of COVID-19 have changed dramatically in a short period of time. Early during the outbreak, Asia was the most affected region, particularly China; however, nowadays the Americas have become the most affected, driven mostly by the United States and Brazil. ^4^ Moreover, mortality has an important variability between highly affected countries. For instance, South Korea has very low mortality rates, exhibiting efficient testing strategies and excellent responses to the emergency. In contrast, countries with less testability, weaker health systems, and poorer overall responses to the virus, reported higher attack mortality and fatality rates^5.^ For this reason and in the absence of a study addressing the epidemiological characteristics of COVID-19 in the DR, this analysis aims to analyze the spatial distribution of COVID-19 and its correlation with the Human Development Index (HDI) in the DR from March 2020 to March 2021.

## Methods

A copy of the COVID -19 Our World in Data database^6^ was obtained in Microsoft Excel format from which data from the DR were extracted. In order to calculate the mortality rate by province, yearly regional and provincial 2000-2030 estimates and projections of the total population according to the National Statistical Office (ONE) were used ^7^. The bulletins of the Ministry of Public Health were also used up to the report no. 367. ^8^ To represent geographical information, confirmed cases were located per province using the corresponding ONE code combined with Microsoft Excel’s 2016 Map Function for cartographic representation. The remaining statistical analyzes were also carried out using solely the Microsoft Excel 2016 spreadsheet. The Human Development Index and its indicators was downloaded from the database published by UNDP (United Nations Development Programme) in 2016 ^9^ and a simple linear regression was performed to predict the incidence and mortality rate from the HDI in Microsoft Excel 2016

## Results and Discussion

### Geotemporal analysis at the national level

During the period March 2020 to March 2021 there were 248,502 cases with 3,262 deaths in total. 1,263,135 tests were performed (88.28 % PCR and 11.72% antigen tests), reporting a daily positivity of 13.47%.

For the accumulated number of cases and deaths from COVID-19, the periods with the highest rates of increase in 2020 occurred between early July and August, reaching its highest peak on September 5 with 2,147 new confirmed cases (Figure 1). Of note, the national elections were originally scheduled for May 17, but were postponed and carried out in July 2020. Although these developed in an organized manner, gatherings and meetings between citizens may explain the increased number of cases. Similar events were observed in Brazil where the highest rates of cumulative cases and deaths from COVID-19 occurred between early July and October 20 2020, during the municipal elections. ^10^

**Figure 1.**
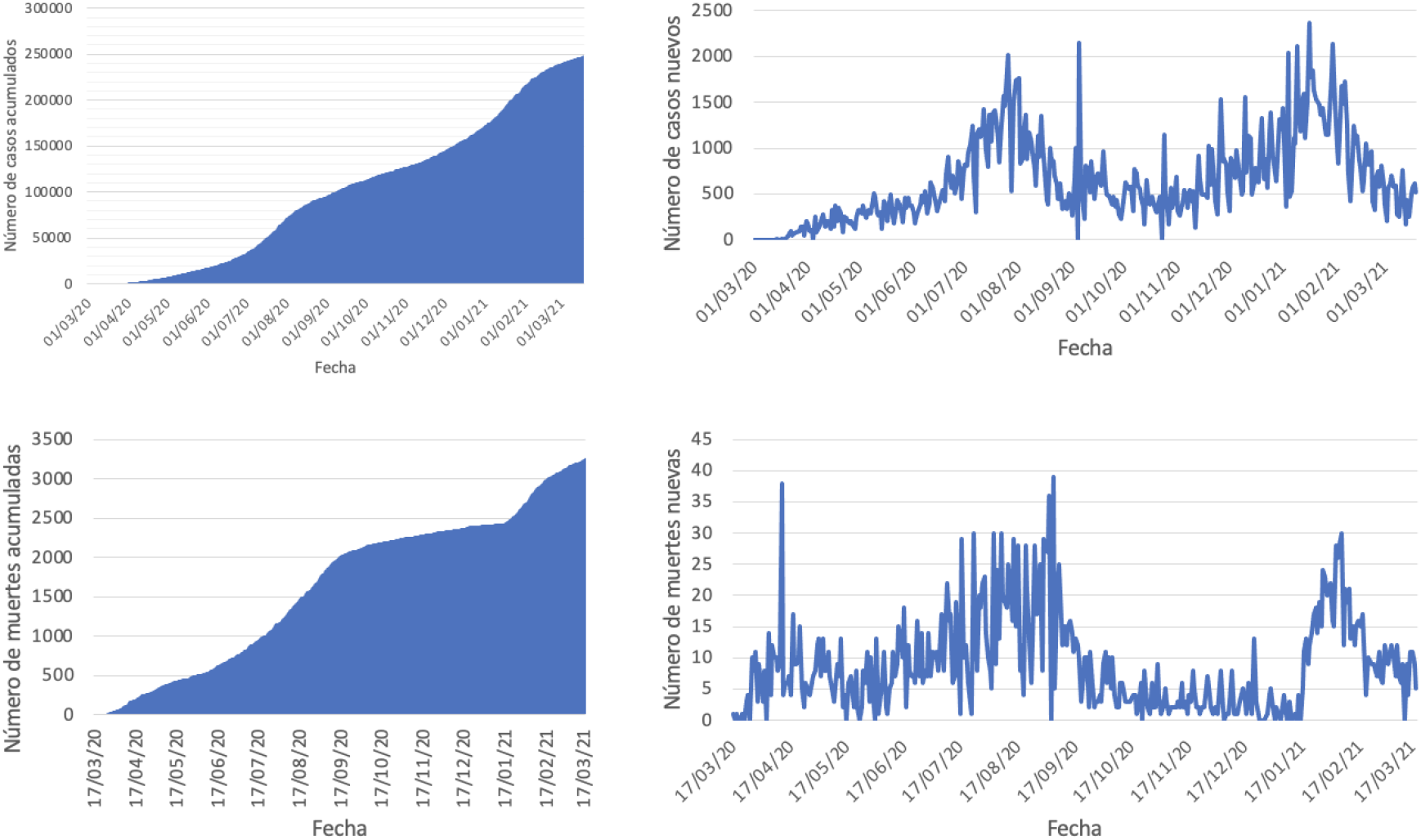
accumulated number of cases and deaths from COVID-19

The maximum number of daily deaths was reported on April 12 of 2020 and September 5 of 2020. As of March 19 of 2021, 3,268 deaths from COVID-19 have been registered in the country (Figure 1). The number of deaths showed a marked decrease between September 9 of 2020 and January 18 of 2021. Duarte province reported the highest mortality rate per 100,000 habitants. A similar geotemporal analysis carried out in Nigeria concluded that Sokoto state had the highest mortality rate (11.11%), suggesting deficient health services. Both the study of Nigeria and ours agreed that low quality healthcare and insufficient testing centers can contribute to increased mortality rates from COVID-19.^11^

Furthermore, the provinces with the highest total number of registered cases were those with the highest population density in the country, including the Distrito Nacional (71,613 cases), Gran Santo Domingo (49,759 cases), and Santiago (27,632 cases). Similarly in the study of Brazil, Campo Grande city, capital of Mato Grosso do Sul, has the highest population density (906,092 habitants) and also registered the highest COVID-19 contamination rates and deaths.^10^ Duarte province had a mortality rate of 68.94 per 100,000 habitants. Some factors that may contribute to the consistent increase in the death rates from COVID-19 in this province include low quality of care, lack of sufficient information on COVID-19 management, and insufficient testing centers.

The 7-day moving average is used to analyze the increasing number of COVID-19 cases and deaths providing an average line over time and is calculated by taking the last 7 days, adding them up, and dividing it by 7 ^12^. In the DR, it showed a significant upward trend for confirmed cases and deaths (Figure 2). The 7-day moving average peak for new cases occurred on July 30 of 2020, while the The 7-day moving average peak for deaths was observed on September 6 of 2020. This was followed by a marked decline that subsequently reached a new peak on February 11 of 2021.

**Figure 2.**
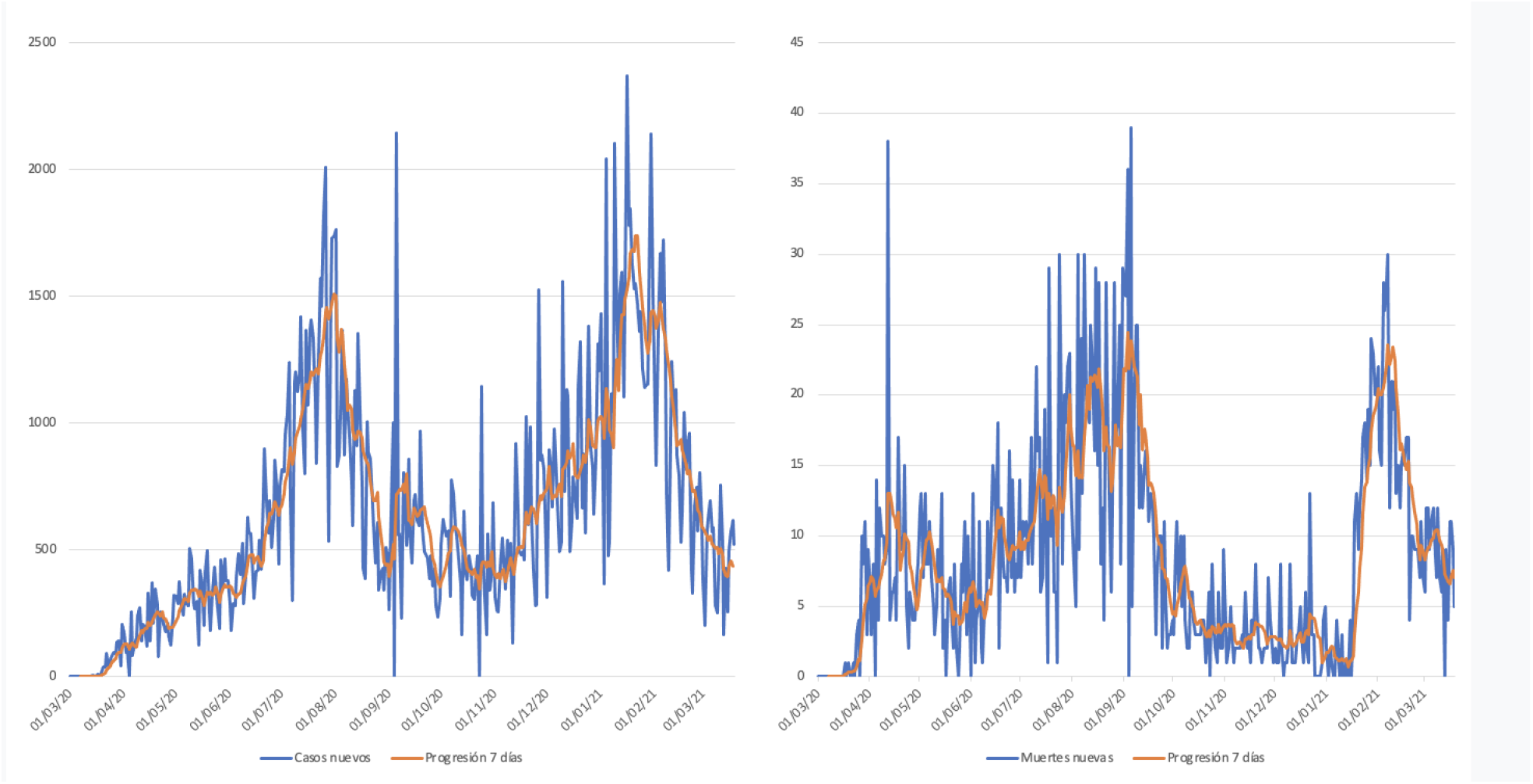
Moving average of the last 7 days for the number of new cases and new deaths from COVID-19 in the Dominican Republic in the period from March, 2020 to March 19, 2021

The positivity rate by month is defined as the percentage of people who test positive among those who have been tested. ^13^ The highest positivity rate (40%) was achieved on August 1, 2020, notably succeeding the general elections held in July. This peak was followed by a relatively constant decrease between August and December of 2020. However, a new peak was observed (30%) in January 2021, right after Christmas holidays. After February 2021 the daily positivity dropped again considerably to 13.47% (Figure 3).

**Figure 3.**
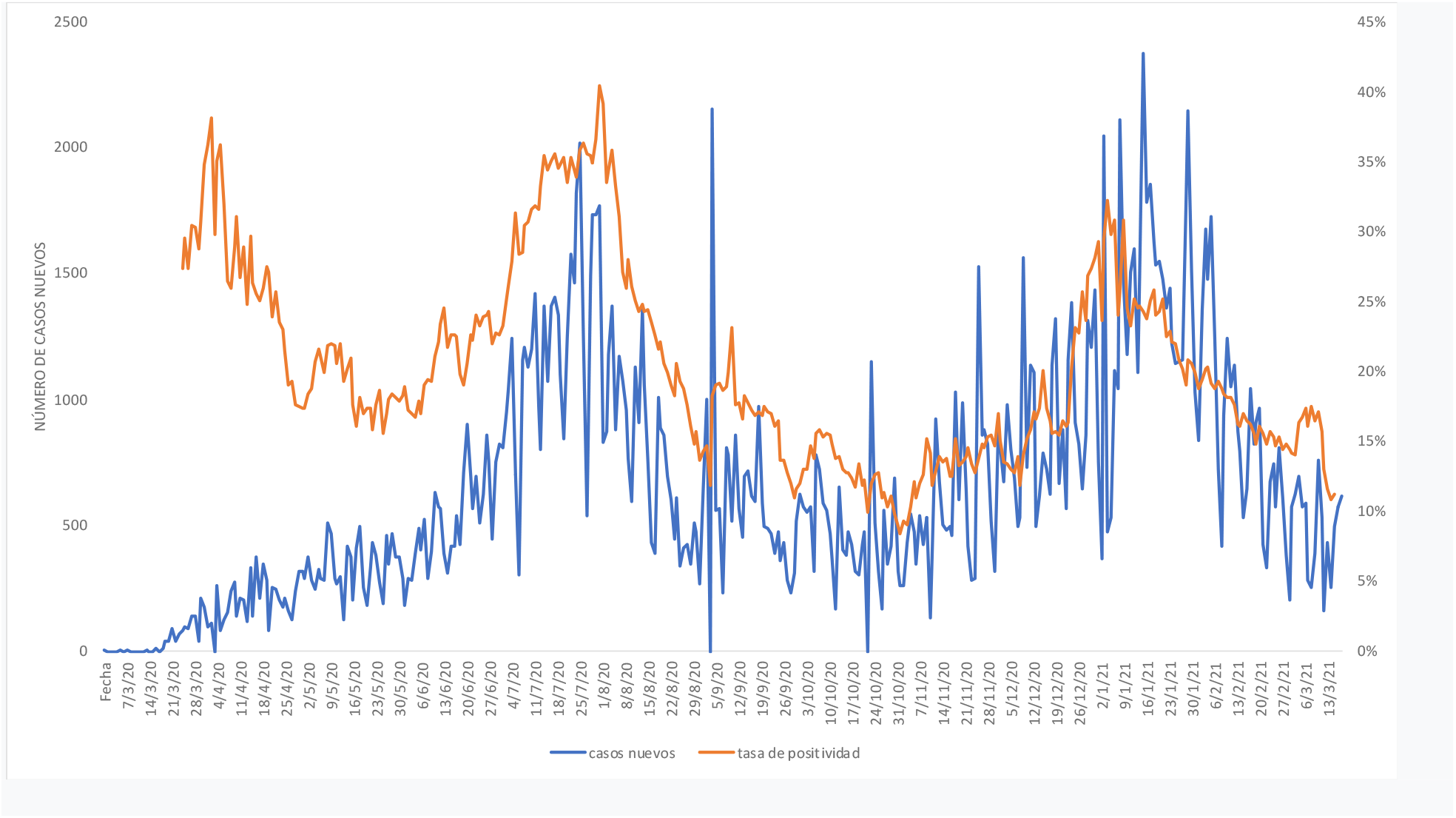
Covid-19 case positivity rate in the Dominican Republic

### Geo-temporal analysis by province

The first confirmed case of COVID-19 in the DR was reported on March 1 of 2020 in the Altagracia Province, since then until March 20, 2021, 249,463 cases of COVID-19 have been reported countrywide. Distrito Nacional (71,613 cases), Santo Domingo (49,759 cases), Santiago (27,632 cases), La Vega (10,047 cases) and La Altagracia (9,193 cases). Among the provinces with the highest number of deaths per 100,000 population are Distrito Nacional (48.49), Santiago (49.39), La Romana (49.43), Duarte (68.94), and Espaillat (48.13).. Figure 4 shows a series of maps with the geotemporal analysis of the status of COVID-19 from March 2020 to March 20, 2021. Of note, the highest rate of new cases was observed on December of 2020, potentially attributed to the Christmas holidays.

**Figure 4.**
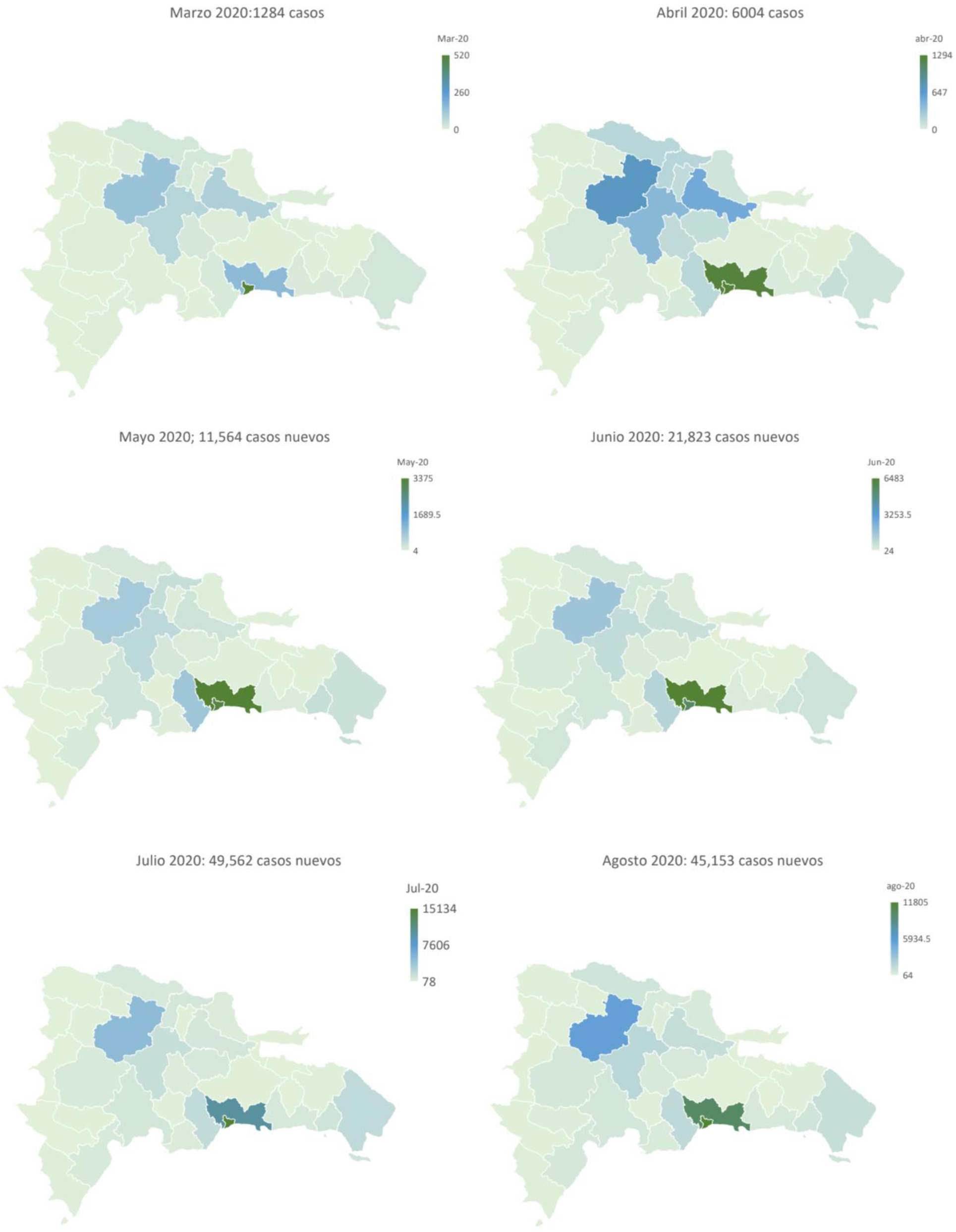
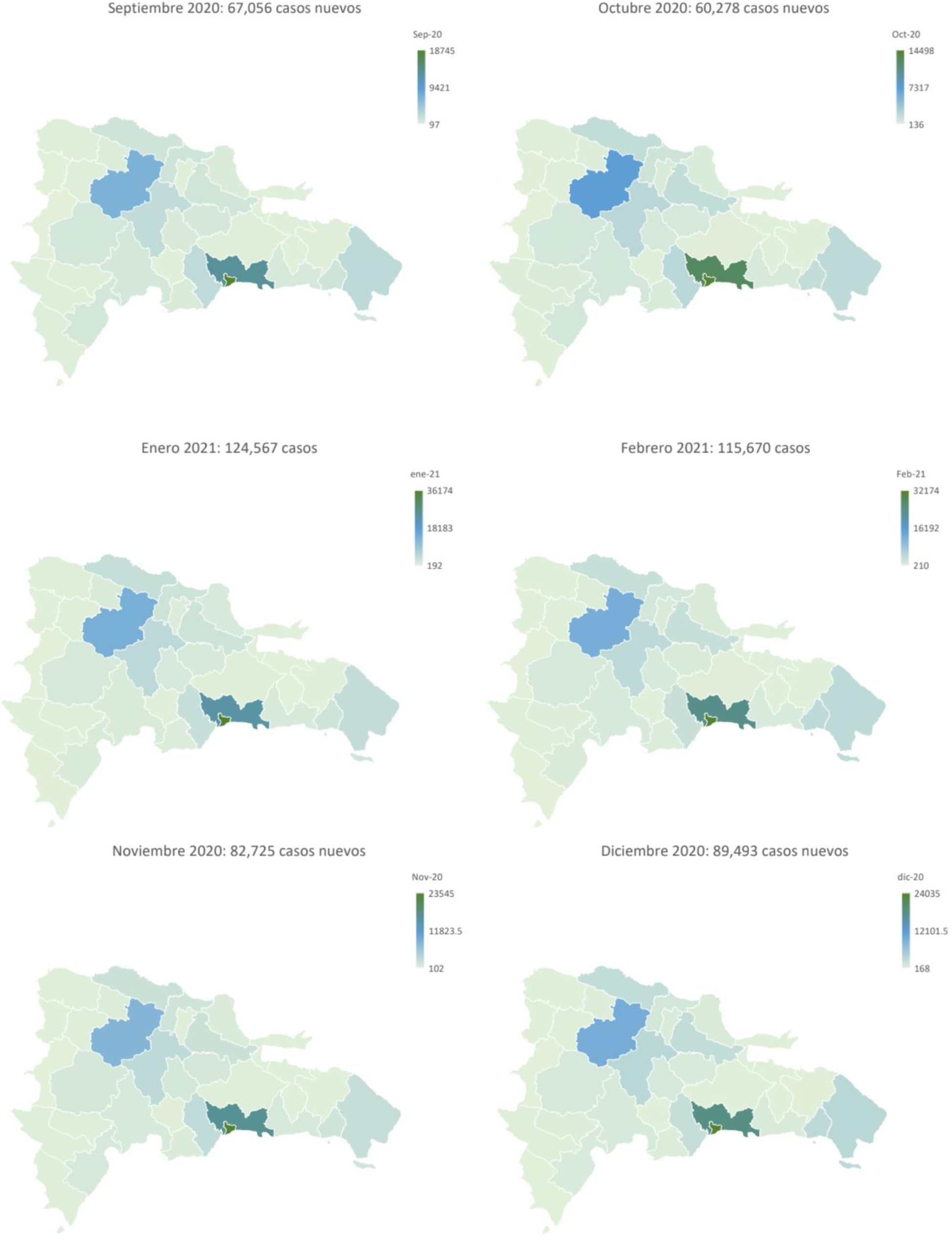
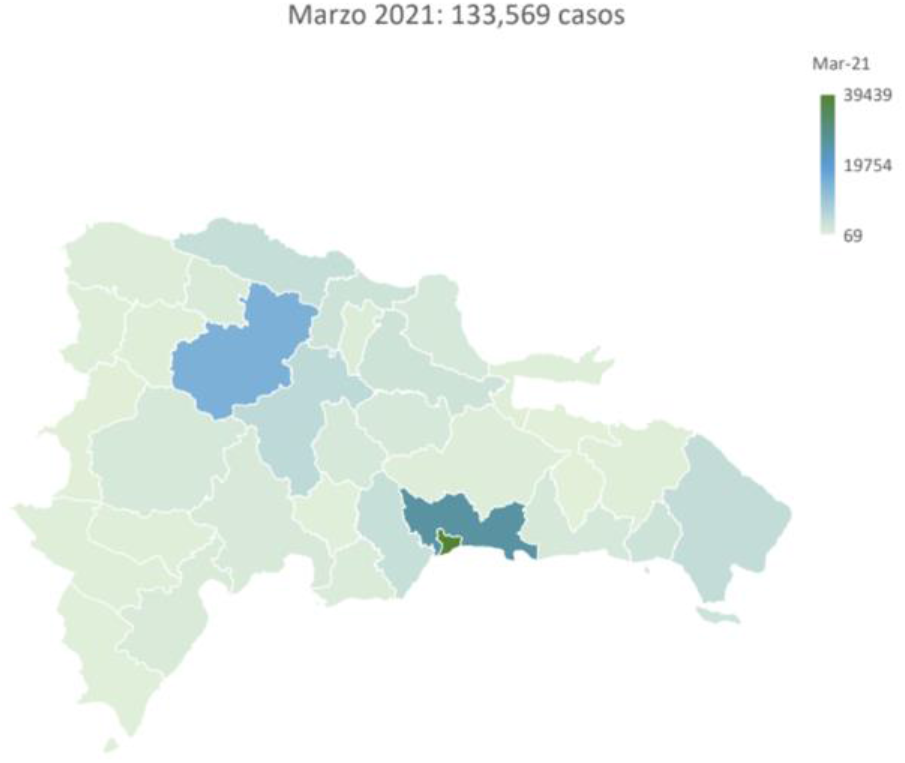
Geotemporal distribution of accumulated cases of COVID-19 in the Dominican Republic between March 1, 2020 and March 20, 2021

### Analysis of the Human Development Index by provinces in the Dominican Republic

The human development index (HDI) explained a significant amount of the variance in the incidence rate, F (1,30) = 33.466, p < 0.00 1, R ^2^ = 0.527, adjusted R ^2^ = 0. 512. The regression coefficient (B = 0. 107, 95% CI [0. 069 -0. 145]) indicated that an increase of 1.0 in HDI corresponded to an increase of 10.7% in the incidence rate per province. A simple linear regression was used to predict death rates from COVID-19 from the Human Development Index. The HDI explained a significant amount of the variance in the mortality rate, F (1.30) = 14.822, p < 0.00 1, R ^2^ = 0. 331, adjusted R ^2^ = 0 .308. Remarkably, the regression coefficient (B = 1,177, 95% CI [55.032 - 179.38]) indicated that an increase of 1.0 in HDI corresponded to an increase of 117% in the mortality rate by province in the country (Figure 5).

**Figure 5.**
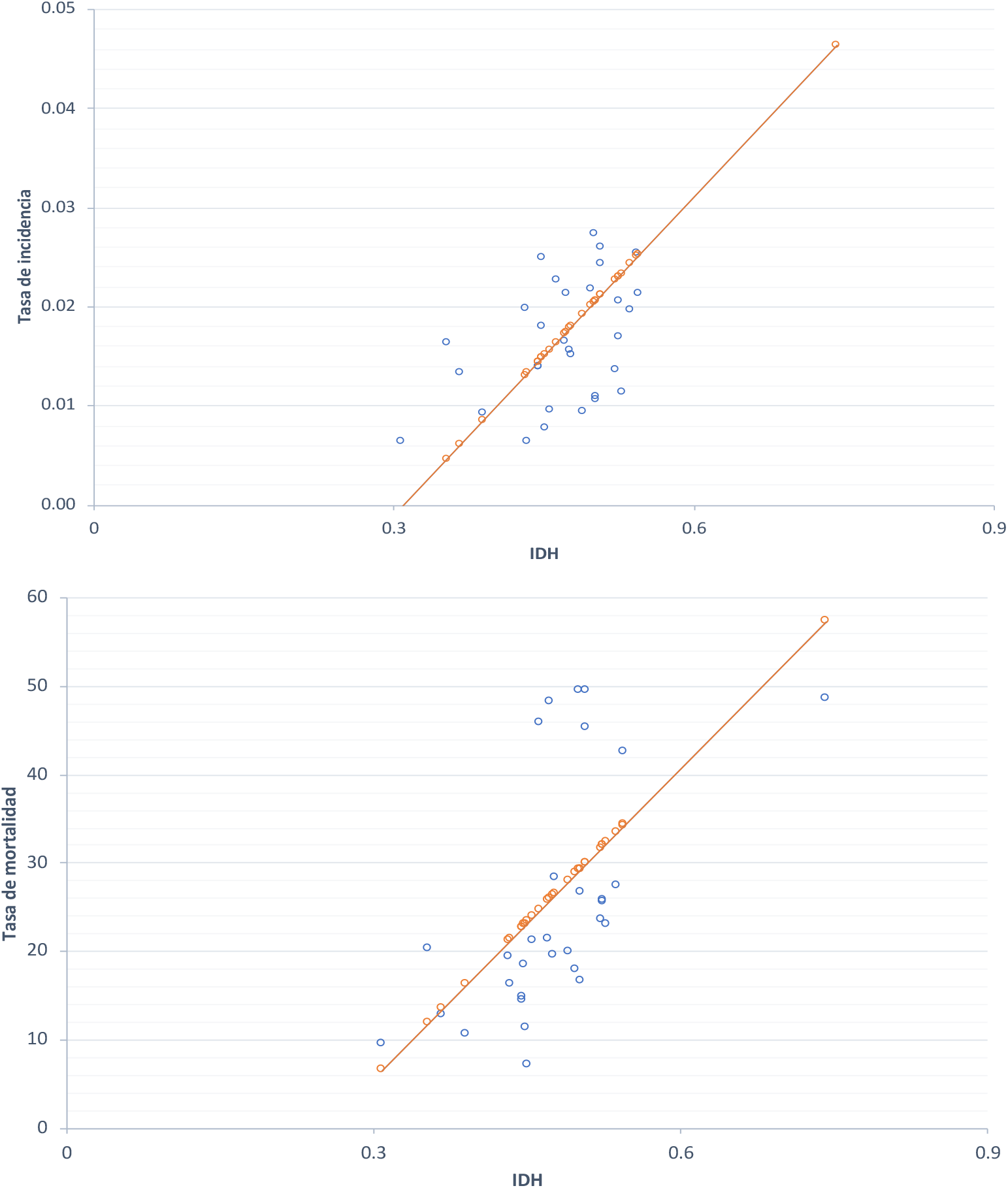
Human development index as a predictor of incidence and mortality rate

Of note, the provinces with the highest human development indexes were those having higher population densities and number of confirmed cases, including Distrito Nacional, Santo Domingo, and Santiago. The fact that a higher HDI predicts a higher incidence rate can be due to areas with lower HDI in the DR having fewer indoor spaces with air conditioning, therefore contributing to disease spreading. Provinces with high HDI have higher longevity rates and hence higher incidence and mortality rates. These higher HDI areas also have greater testing services and access to medical care, for this reason, the population living in the provinces with fewer resources migrate to these areas seeking medical care, which can account for the relation between higher HDIs and higher mortality rates.

In Brazil, low HDI revealed population vulnerability and gaps in access to health services, as denoted by the example of the City of Campo Grande having one of the higher incidence and mortality rates from COVID-19. ^11^ Comparably in Italy, high HDI rates also suggest that developed regions have the highest Covid-19 cases. ^14^

To further explore how HDI values are associated with the incidence rate, GDP data by provinces in the DR for 2016 was obtained, where a positive correlation between per capita income in DOP and the incidence rate was observed with a value of r (31) = 0.83, p <0.001.

## Conclusion

This article evaluated the spatial effects of the COVID - 19 pandemic at the provincial and national levels. Provinces with the highest incidence and mortality rates were analyzed from March 2020 to March 2021 and compared with their corresponding HDI. We believe that the geo - temporal analysis combined with the Human Development Index can contribute to developing responses to face the pandemic in the country. The results obtained in this study contribute to the knowledge on the epidemiological development of the virus in the DR and thus, open new lines for future analysis on COVID-19. It is also concluded that extensive prevention strategies should continue to be strengthened to reduce the contagion curve, lower the disease spread and diminish its impact on the HDI. This analysis leads to valuable information on the dynamics of COVID-19 in the country that can serve as a guide for implementing new measures at the national and local level. the lack of information in government sources was a limitation for this study

## Data Availability

6. Ritchie, Hannah; Ortiz-Ospina, Esteban; Beltekian, Diana; Mathieu, Edouard; Hasell, Joel; McDonald, Bobbie; Giattino, Charlie; Appel, Max; Roser, Max. (2020). Dominican Republic: Coronavirus Pandemic Country Profile. Retrieved on April 20, 2021, from Our World in Data: https://ourworldindata.org/coronavirus/country/dominican-republic 7. National Statistics Office. Provincial projections and estimates 2000-2030. Dominican Government: ONE; 2016 [accessed 03/13/2021]. Available at: https://www.one.gob.do/demograficas/proyecciones-de-poblacion 8. Ministry of Public Health. COVID- 19. (April 8, 2020). Recovered 20 Marzo 2021 of Gob.do website : https://www.msp.gob.do/web/?page_id=6948

https://www.one.gob.do/demograficas/proyecciones-de-poblacion

https://www.msp.gob.do/web/?page_id=6948

## References

1. Khan M, Khan H, Khan S, Nawaz M. Epidemiological and clinical characteristics of coronavirus disease (COVID-19) cases at a screening clinic during the early outbreak period: a single-center study. J Med Microbiol. 2020 Aug; 69 (8): 1114–1123. doi: 10.1099/jmm.0.001231. PMID: 32783802; PMCID: PMC7642977

2. Zhou, M., Zhang, X., & Qu, J. (2020). Coronavirus disease 2019 (COVID-19): a clinical update. Frontiers of medicine, 14 (2), 126–135. https://doi.org/10.1007/s11684-020-0767-8

3. World Health Organization. Coronavirus disease 2019 (COVID-19) Situation Report - 38. Geneva: WHO; 2021 [accessed 03/19/2021]. Available at https://covid19.who.int/table?tableChartType=heat

4. Wiersinga WJ, Rhodes A, Cheng AC, Peacock SJ, Prescott HC. Pathophysiology, Transmission, Diagnosis, and Treatment of Coronavirus Disease 2019 (COVID-19): A Review. JAMA. 2020 Aug 25; 324 (8): 782–793. doi: 10.1001/jama.2020.12839. PMID: 32648899.

5. Ortiz-Prado, E., Simbaña-Rivera, K., Gómez-Barreno, L., Rubio-Neira, M., Guaman, LP, Kyriakidis, NC, Muslin, C., Jaramillo, A., Barba-Ostria, C., Cevallos-Robalino, D., Sanches-SanMiguel, H., Unigarro, L., Zalakeviciute, R., Gadian, N., & López-Cortés, A. (2020). Clinical, molecular, and epidemiological characterization of the SARS-CoV-2 virus and the Coronavirus Disease 2019 (COVID-19), a comprehensive literature review. Diagnostic microbiology and infectious disease, 98 (1), 115094. https://doi.org/10.1016/j.diagmicrobio.2020.115094

6. Ritchie, Hannah; Ortiz-Ospina, Esteban; Beltekian, Diana; Mathieu, Edouard; Hasell, Joel; McDonald, Bobbie; Giattino, Charlie; Appel, Max; Roser, Max. (2020). Dominican Republic: Coronavirus Pandemic Country Profile. Retrieved on April 20, 2021, from Our World in Data: https://ourworldindata.org/coronavirus/country/dominican-republic

7. National Statistics Office. Provincial projections and estimates 2000-2030. Dominican Government: ONE; 2016 [accessed 03/13/2021]. Available at: https://www.one.gob.do/demograficas/proyecciones-de-poblacion

8. Ministry of Public Health. COVID-19. (April 8, 2020). Recovered 20 Marzo 2021 of Gob.do website : https://www.msp.gob.do/web/?page_id=6948

9. UNDP. (2017). Human Development Report 2016: Human Development for All People. New York, NY: United Nations.

10. De Souza, A., Abreu, MC, & de Oliveira-Júnior, JF (2021). Spatio -temporal analysis between the incidence of COVID-19 and human development in Mato Grosso do Sul, Brazil. doi: 10.1101/2021.01.19.21250106

11. Suleiman, AA, Suleiman, A., Abdullahi, UA, & Suleiman, SA (2021). Estimation of the case fatality rate of COVID-19 epidemiological data in Nigeria using statistical regression analysis. Biosafety and health, 3 (1), 4–7. https://doi.org/10.1016/j.bsheal.2020.09.003

12. Georgia Rural Health Innovation Center. What is a moving average, and why is it useful?. [accessed 6/11/21]. Available at Georgiaruralhealth.org website: https://www.georgiaruralhealth.org/blog/what-is-a-moving-average-and-why-is-it-useful/

13. Castillo, A. (May 8, 2020). This is how the positivity rate is calculated in a pandemic. Free Journal. Retrieved April 11, 2021, from https://www.diariolibre.com/actualidad/salud/asi-se-calcula-la-tasa-de-positividad-en-una-pandemia-EM18726750

14. Ghosh, P., & Cartone, A. (2020). A Spatio - temporal analysis of COVID - 19 outbreak in Italy. Regional Science Policy & Practice, 12 (6), 1047–1062. https://doi.org/10.1111/rsp3.12376

